# Low cost local food supplements could improve maternal and birth outcomes: a pilot randomized controlled trial

**DOI:** 10.1101/2020.08.24.20136762

**Authors:** Kun A Susiloretni, Dyah N Subandriani, Elisa Ulfiana, Trina Astuti, Sunarto, Emily R Smith

## Abstract

**Objectives:** Maternal nutrition plays a crucial role in influencing fetal growth and birth outcomes. The government of Indonesia has provided a manufactured fortified food supplementation program for undernourished pregnant mothers. We hypothesized a low-cost local food supplementation (LFS) would improve maternal and birth outcomes compared with government food supplementation (GFS).

**Methods:** We conducted a randomized control trial. We enrolled 54 pregnant mothers with MUAC<23.5cm who were assigned into two groups. The intervention group received LFS and multiple micronutrient supplements (MMS). The control group received the GFS and iron and folic acid supplements (IFA). LFS was a balanced energy protein food supplement made from nuts and fish. LFS and GFS contained approximately 500kcal energy and 10grams protein. We compared the effectiveness of these two interventions using logistic and linear regression.

**Results:** At the baseline, the two groups had similar characteristics. After 60 days of treatment, mothers in the LFS group were 2.28 folds more likely to increase MUAC (RR 2.28; 95%CI 1.58,3.27, p<0.001) and 4.73 folds to increase gestational weight (RR 4.73; 95%CI 1.37,16.3, p=0.015) than were mothers in the GFS group. For birth outcomes, in the LFS group had reduction of birthweight <3000grams (RR 0.15; 95%CI 0.023,0.98; p=0.048), short birth length (RR 0.068; 95%CI 0.005,0.93; p=0.044), and cesarean delivery (RR 0.11; 95%CI 0.022,0.61, p=011) as compared to the GFS group.

**Conclusions for Practice:** Local foods and MMS supplementation can improve maternal and birth outcomes. Therefore, local food supplements can be considered for food supplementation programs to undernourished pregnant mothers.

**Significance Statement:** *What is already known on this subject?:* Balanced protein-energy foods fortified with multiple micronutrients supplements has been known as an effective intervention to improve birth length and birth weight for underweight pregnant women.

*What this study adds?:* The provision of balanced energy protein of local foods supplements combined with multiple micronutrients supplements pills, showed the reduction on birth weight < 3000grams and short birth length. Surprisingly could reduce cesarean deliveries, and furthermore could give a chance to produce local food supplements by micro and small enterprises in community setting. Future studies should be conducted using better research design and embeded in the food and stunting reduction system.

## Introduction

Maternal nutrition plays a crucial role in influencing fetal growth and birth outcomes. A prospective cohort study in Ethiopia showed that mothers who consumed ≥ 4 food groups had a reduced risk of maternal anemia, preterm delivery, and low birth weight (Zerfu, Umeta, & Baye, 2016). Maternal undernutrition contributes to fetal growth restriction, which increases the risk of neonatal deaths and, for survivors, of stunting by 2 years of age (Black et al., 2013). Chronic energy malnutrition mothers with (mid-upper arm circumference (MUAC) < 23.5cm) is a key risk factor for delivering babies with low birth weight, small-for-gestational age, and preterm birth (Sebayang et al., 2012).

Furthermore, maternal nutrition predicts health and wellness in later life. A mother’s diet during pregnancy may affect fetal programming the baby (Godfrey & Barker, 2001). During development in the utero, there are critical periods during which organs and systems mature. Malnutrition and other adverse influences during development may permanently alter gene expression. They also lead to a slowing of growth, which is associated with increased vulnerability to chronic diseases. They can reduce key organ’s function, altered settings in metabolism and hormonal feedback, and more vulnerability to adverse environmental influences in later life (D. J. Barker, 2012; D. J. P. Barker, 2007; Branca, Piwoz, Schultink, & Sullivan, 2015; Godfrey & Barker, 2001; Warner & Ozanne, 2010).

However, inadequate maternal nutrition and related complications continue to be common in Indonesia. In 2018, the maternal mortality rate was 305/100,000 live births and the infant mortality rate was 34 /1,000 live births, while baby with low birth weight was 6.2%, short birth length was 22.6%, and preterm birth was 29.5%. The prevalence of chronic energy malnutrition in pregnant mothers (mid-upper arm circumference (MUAC) <23.5cm) was 17.3% and nearly half of all pregnant mothers were anemic. (Ministry of Health Republic of Indonesia, 2018). The national dietary survey reported that pregnant mothers consumed protein less than 80% recommendations were 49.6% and consumed energy less than 70% recommendation was 52.9% (National Health Institute of Research and Development, 2014).

The government of Indonesia has provided the fortified food supplementation (GFS) with multiple micronutrient supplements (MMS) for undernourished pregnant mothers. The GFS has been distributed across the nation from a single manufacture. The consequence is the high cost for distribution to 44 provinces. To minimize the cost and support for a healthy diet from the sustainable food system (Willett et al., 2019), it is needed to provide local food supplementation that can consume combining with MMS. The most recent study, MMS supplementation for pregnant mothers could improve survival for female neonates and provided greater birth-outcome benefits for infants born to undernourished and anemic pregnant women (Haider & Bhutta, 2017; Smith et al., 2017). While balanced energy protein supplementation provided greater benefit on birth weight by 59.89 g and reduction of small for gestational age by 31% compared to the control group (Imdad & Bhutta, 2011).

This study aimed to assess whether a low-cost local food balance energy protein supplementations (LFS) and MMS could improve maternal and birth outcomes compared with government fortified food supplementation (GFS) and iron-folic acid (IFA).

## Methods

### Trial design

We considered this study as a randomized, open-label, pilot study (Eldridge et al., 2016) to investigate feasibility of the LFS for pregnant women intervention. This pilot trial will help to inform sample size requirements for a future definitive RCT. Pregnant women were randomly allocated in a 1:1. This study was registered at https://clinicaltrials.gov as NCT02959125.

### Participants

Sixty pregnant mothers were recruited. They met the following inclusion criteria: pregnant mothers who had visited village midwives for antenatal care, with single pregnancies, gestational age at less than 28 weeks, had MUAC <23.5cm, singleton, and had no intention of moving from the area for at least 6 months. We excluded mothers who had a current stillbirth or miscarriage to avoid having the follow-up uncompleted. These rigid criteria would limit generalizability.

The study took place in the Demak District, Central Java Province, Indonesia from September 2016 to August 2017. We selected the Karangawen2 Public Health Center area because it has the second-highest chronic energy malnutrition (MUAC<23.5cm) prevalence in the Demak District (11.8%). This administrative area covers 36.4 km^2^ and includes 6 villages. The total population is 48,288 people. The population density is 1,327 people/km^2^. There were 992 pregnant mothers in the study area. The main occupations for people in the study area included: farmers, farmworkers, construction workers, industrial workers, and traders. The coverage of complete antenatal care visits was 95%, and all births were attended by a skilled birth attendant.

The study was approved by the Semarang Health Polytechnic research ethics review committee. Demak District Licensing Service Agency and Karangawen Public Health Center also granted the approvals. Written informed consents were obtained from all participants.

### Interventions

We developed balanced energy protein food supplementations (LFS) using protein source foods that are cheap and made from locally available foods including fish, peanut, green bean, soybean, and brown bean. Beans are also a source of micronutrients needed by pregnant, including folate, magnesium, potassium, and zinc (Messina, 2014). We chose cookies as the form of food supplementations because it is a familiar food, easy to cook, and had a relatively long shelf life because of low moisture content. The five cookies types were produced at the Culinary Laboratory at the Semarang Health Polytechnic. The sensory evaluation was carried out using four-scale hedonic test to the participants, yielded that LFS had better score ranging from 3.10 to 3.51 compared to 2.96 to 3.24 for GFS biscuits.

In this study, all participants received a one-day class on maternal health and early life nutrition. The trainers were from Public Health Centers and researchers. The trainers were provided with power-point presentation slides, while participants received a hand out of power-point presentation slides. After training, we gave food and nutrition supplementation package for 30 days. The treatment group received the LFS (i.e. balanced energy protein food supplementation), which contained approximately 512 kcal and 11.4 grams protein per package/day and MMS pills. The control group had received the existing government fortified food (GFS) contained approximately ~485 kcal and ~15.8 grams protein per package/day and also iron-folic acid (IFA) supplementations. The GFS is a cookie made from wheat flour, egg and dried egg, milk, butter and sugar which fortified with multiple micronutrients.

Multiple micronutrient composition of supplement or fortification used in this study compare to other studies is shown in Table 1. The duration of the intervention was 60 days. Macronutrient composition of food supplements can be seen in Table 2, while cookies ingredients show in supplemental file #1.

**Table 1.** Multiple micronutrient composition of supplement used to compare to UNIMMAP

| Micronutrient | LFS<sup>^</sup> | GFS<sup>^</sup> | UNIMMAP | Mexico* | Nepal** |
| --- | --- | --- | --- | --- | --- |
| Iron (mg) | 27 | 15 | 30 | 62.4 | 60 |
| Zinc (mg) | 15 | 7.5 | 15 | 12.9 | 30 |
| Copper (mg) | 20 | - | 20 | — | 2.0 |
| Selenium (µg) | 25 | 35 | 65 | — | — |
| Magnesium (mg) | 100 | - | — | 252 | 100 |
| Iodine (µg) | 150 | 100 | 150 | — | — |
| Vitamin A (IU) | 5000 | 800 | 800 µg | 2,150 | 1,000 |
| β-Carotene (mg) | - | - | — | — | — |
| Vitamin B1 (mg) | 10 | 1.3 | 14 | 0.93 | 1.6 |
| Vitamin B2 (mg) | 10 | 1.4 | 14 | 1.87 | 1.8 |
| Folic acid (µg) | 400 | 600 | 400 | 215 | 400 |
| Niacin (mg) | 20 | 18 | 18 | 15.5 | 20 |
| Vitamin B6 (mg) | 10 | 17 | 19 | 1.94 | 2.2 |
| Vitamin B12 (µg) | 30 | 26 | 26 | 2.04 | 2.6 |
| Vitamin C (mg) | 90 | 85 | 70 | 66.5 | 100 |
| Vitamin D (µg) | 400 | 5 | 5 | 309 | 10 |
| Vitamin E (IU) | 30 | 15 | 10 | 5.7 | 10 |
| Vitamin K (µg) | — | - | — | — | 65 |
| Calcium (mg) | 162 | 250 | — | — | — |
| Biotin mcg | 45 | — | — | — | — |
| Phosphorus (mg) | 125 | — | — | — | — |

<sup>^</sup> Macronutrient composition based on information from the label of nutrition facts panel.
\*Mexico (Ramakrishnan, González-Cossío, Neufeld, Rivera, & Martorell, 2003)
\*\*Nepal (Christian et al., 2003)

**Table 2.** Macronutrient composition of LFS and GFS

| Nutrients | FFS | **GFS** |
| --- | --- | --- |
| Moisture | 3.64 | 4.20 |
| Ash | 1.55 | 1.33 |
| Protein | 11.42 | 12.98 |
| Fat | 24.35 | 29.10 |
| Carbohydrate | 59.04 | 34.55 |
| Energy | 512.41 | 465 |

Note: Macronutrient composition was determined from the mix of cookies which measured by the Local Health Laboratory of Semarang.

### Outcomes

The primary outcome was infant weight at birth. The secondary outcomes were body weight and MUAC increment of pregnant mothers, birth length, preterm birth, and delivery method. Maternal outcomes were MUAC and body weight increment, assessed approximately 65-77 days after initiation of food supplementation. Because of the time difference of these measurements, we used average of MUAC increment and body weight gain within 30days. MUAC was measured to the nearest millimeter using numeral insertion tapes. The weight of pregnant mothers was measured to the nearest 0.1kg using a digital scale.

Birth outcomes were assessed by research staff during home visits after delivery. Birth outcomes data (gestational age at birth, delivery method, birth weight, and length) were obtained from the ‘maternal and child health book record’ belonging to the mothers. The books were filled by health professionals where the mothers giving birth at a health facility. Gestational age was calculated from the reported first day of the last menstrual period and prospectively collected histories of menstruation. Preterm delivery was defined as delivery before 37 weeks of gestation. They were then categorized into binary variables: term and preterm, vaginal and cesarean delivery methods, birthweight using cut-off point 3000grams, and short birth length using cut-off point 48cm.

At the baseline, we measured energy and protein dietary intake, maternal and paternal age, maternal and paternal education, maternal height, parity, antenatal care received, and house owner by home visits. Dietary intake was assessed by 2 x 24-h dietary recall, then converted to the nutrient adequacy intake using Indonesian Food Nutri Survey software. The other measurements were assessed by structured interviews with questionnaires (Osendarp et al., 2000). All anthropometric measurements, dietary intake, and other assessments were performed by six enumerators who recently completed their bachelor degrees of applied nutrition and received a one-day training.

### Sample size

Since this was a feasibility study for the clinical trial phase, a sample size calculation was not performed. We recruited 60 pregnant mothers, as the main aim was to obtain a quantitative measure of the standard deviation of effect size to inform the sample size calculation for a future RCT (Eldridge et al., 2016).

### Statistical analysis

We tested the homogeneity baseline characteristics using Chi-Square or Fisher Exact test, independent t-test, or rank Spearman test. Multivariable linear and logistic regressions were used to assess effectiveness. For birth weight, birth length, delivery methods, and gestational age at birth, we used logistic regressions adjusted by sex of the child, parity, maternal height and MUAC at birth, maternal age, maternal education, and house owner. While for gestational weight and MUAC increment we used linear regression adjusted by maternal MUAC and body weight at baseline, food supplements (GFS or LFS) consumed, and micronutrient supplementation consumed, maternal age, gestational age at enrollment, parity, maternal height, protein and energy adequacies. All analysis was completed using STATA 14.

## Results

Figure 1 shows the flow of participants through the trial. We recruited 76 pregnant women from village midwife records indicating MUAC <23.5cm, and they were randomized to the LFS and GFS group. Women attended the training and were checked for eligibility. We found 30 mothers in the LFS group and 17 mothers in the GFS group had a confirmed MUAC <23.5cm and give consented to participate in the study. To fulfill a minimum sample size, we looked for 13 mothers consecutively selected from midwives’ record to assign to the GFS group. Finally, at the baseline, we could assign 30 mothers in the LFS and 30 mothers in the GFS group. After 60 days of treatment, there were 29 mothers in the LFS group and 25 mothers in the GFS group completed for measuring maternal outcomes. For birth outcomes, there were 25 mothers in the LFS group and 24 mothers in the GFS group completed in this study.

**Figure 1.**
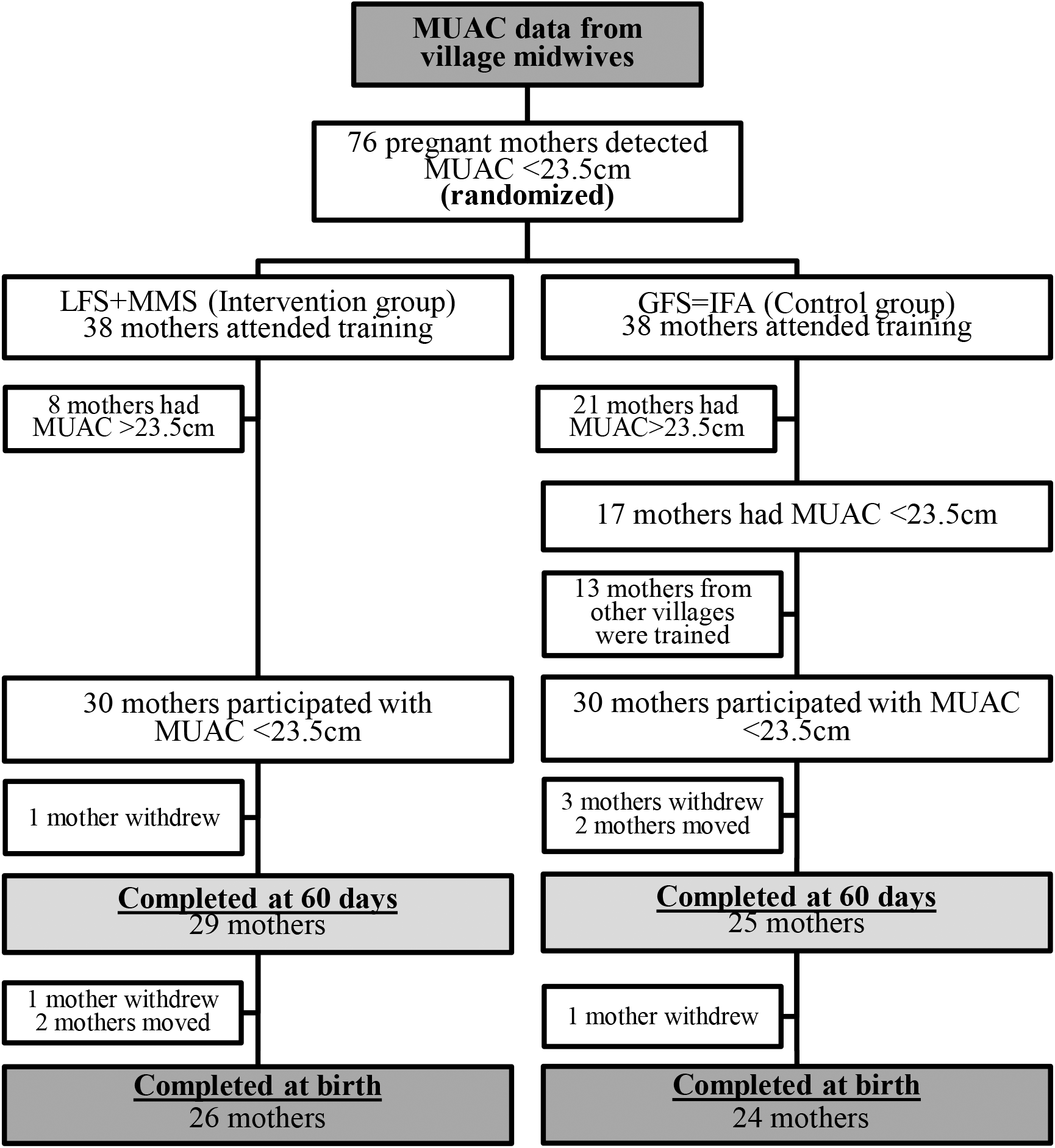
Participants flow diagram of the trial for two study groups.

Participants’ characteristics were similar between LFS and GFS groups, including maternal age, gestational age, parity, height, ANC visits, education, occupation, MUAC, energy and protein intakes, and also paternal age, education, occupation and house owner (Table 3). After 60 days of intervention (Table 4), the mean increment of weight and MUAC per 30 days in the LFS group were 2.25+1.7 kg and 0.43+0.35 cm respectively compared with 2.03+1.1 kg and 0.25 +0.40 cm in the GFS group.

**Table 3.**
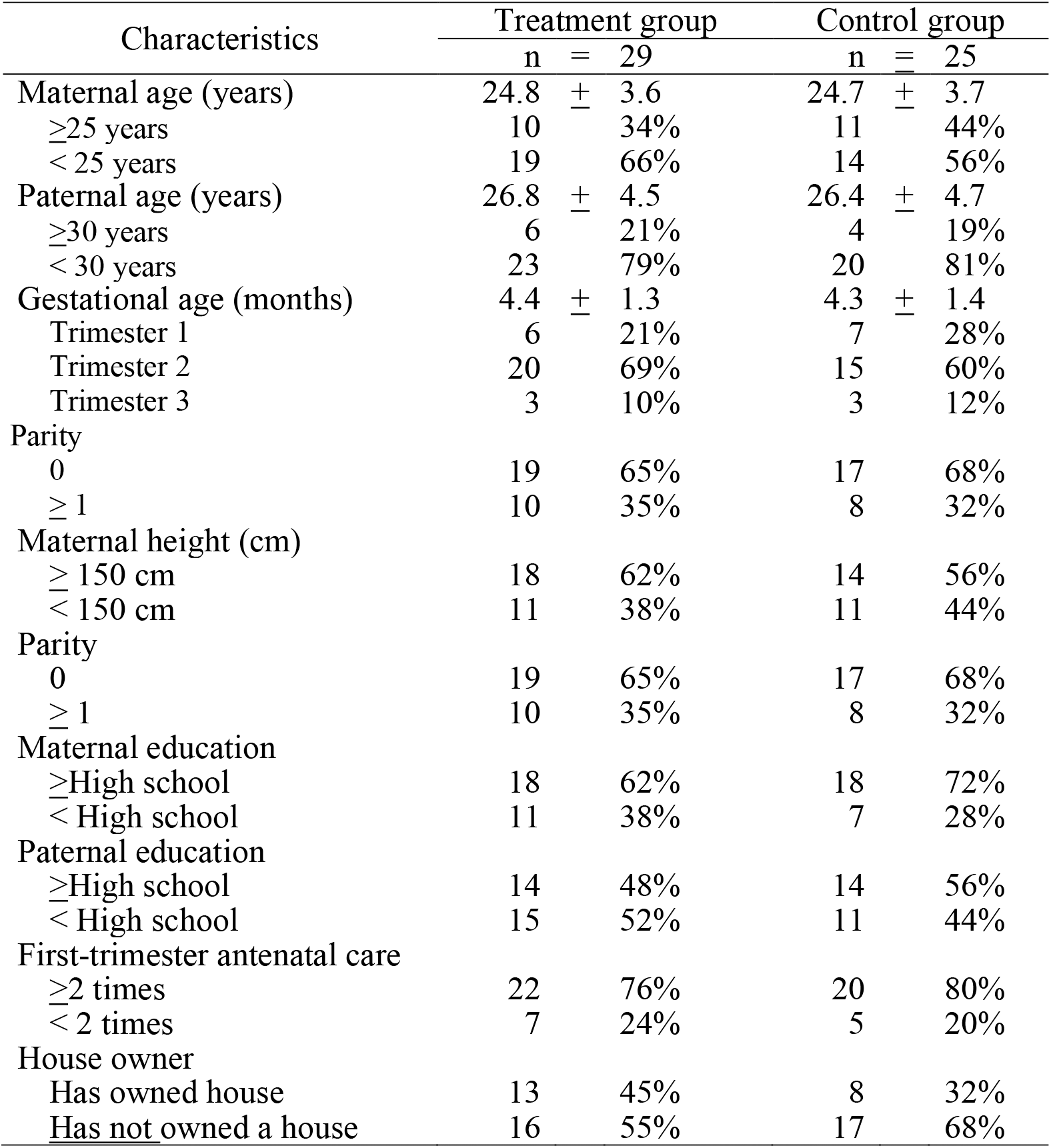
Baseline characteristics at the treatment and control group

**Table 4.** Maternal and birth outcomes after 60 days of treatment and birth outcomes

| Outcomes | LFS + MMS (n=29) |  | GFS + IFA (n=25) |  | p |
| --- | --- | --- | --- | --- | --- |
|  | Mean | ± SD | Mean | ± SD |  |
|  | n | % | n | % |  |
| Maternal outcomes |  |  |  |  |  |
| Gestational weight (kg) |  |  |  |  |  |
| Baseline | 44.6 | ± 4.4 | 44.0 | ± 4.6 | 0.327 |
| Endline | 50.1 | ± 4.8 | 49.0 | ± 5.0 | 0.212 |
| Gestational weight increment (kg)/ month | 2.25 | ± 1.7 | 2.03 | ± 1.4 | 0.132 |
| Mean difference (95% CI) |  | 0.49 [-0.38, 1.37] |  |  |  |
| MUAC (cm) |  |  |  |  |  |
| Baseline | 21.9 | ± 1.3 | 22.0 | ± 1.5 | 0.322 |
| Endline | 23.0 | ± 1.1 | 22.6 | ± 1.5 | 0.107 |
| MUAC increment (cm) / month | 0.43 | ± 0.35 | 0.25 | ± 0.40 | 0.042 |
| Mean difference (95% CI) |  | 0.18 [-0.25, 0.38] |  |  |  |
| MUAC at endline |  |  |  |  |  |
| > 23.5 cm | 7 | 27% | 5 | 22% | 0.674 |
| ≤ 23.5 cm | 19 | 73% | 18 | 78% |  |
| Energy adequacy (%) |  |  |  |  |  |
| Baseline | 81.5 | ± 20.4 | 85.1 | ± 26.7 | 0.713 |
| Endline | 84.2 | ± 26.3 | 83.3 | ± 28.9 | 0.453 |
| Energy intake increased | 2.7 | ± 23.2 | 0.1 | ± 21.2 | 0.334 |
| Protein adequacy (%) |  |  |  |  |  |
| Baseline | 85.6 | ± 32.2 | 85.7 | ± 32.2 | 0.497 |
| Endline | 94.3 | ± 35.5 | 79.1 | ± 28.3 | 0.054 |
| Protein intake increased | 8.0 | ± 32.1 | -5.6 | ± 27.7 | 0.057 |
| Micronutrient received/month |  |  |  |  |  |
| >15 tablets | 15 | 52% | 4 | 16% | 0.006 |
| ≤15 tablets | 14 | 48% | 21 | 84% |  |
| Food supplementation received/month |  |  |  |  |  |
| 15-day packages | 14 | 48% | 8 | 32% | 0.225 |
| 15-day packages | 15 | 52% | 17 | 68% |  |
| Birth outcomes |  |  |  |  |  |
|  | n = 26 |  | n = 24 |  |  |
| Birth weight (kg) | 3142 | ± 236 | 3043 | ± 424 | 0.155 |
| ≥ 3000 grams | 20 | 77% | 13 | 55% | 0.090 |
| < 3000 grams | 6 | 23% | 11 | 45% |  |
| Reduction of birth weight <3000grams |  |  | 22% |  |  |
| Delivery methods |  |  |  |  |  |
| Vaginal | 23 | 88% | 11 | 45% | 0.001 |
| Cesarean | 3 | 12% | 13 | 55% |  |
| Reduction of Cesarean |  |  | 32% |  |  |
| Gestational age at birth (weeks) | 37.9 | ± 1.7 | 37.9 | ± 1.5 |  |
| Term | 18 | 69% | 16 | 67% | 0.846 |
| Preterm | 8 | 31% | 8 | 33% |  |
| Reduction of preterm birth |  |  | 2% |  |  |
|  | n = 24 |  | n = 23 |  |  |
| Birth length (cm) | (48.4 | + 2.2) | (48.1 | ± 2.1) | 0.299 |
| ≥ 48 cm | 22 | 95% | 15 | 70% | 0.027 |
| < 48 cm | 2 | 5% | 8 | 30% |  |
| Reduction of short birth length |  |  | 25% |  |  |

P-value from independent t-test or chi-square – Fisher exact test

Adherence to food and nutrition supplementations in the LFS group was higher compared to the GFS group. The proportion of mothers in the LFS group who reported taking more than 15 pills of nutrient supplementation in a month was 52% in the LFS group compared to 16% in the GFS group (p=0.006). While the proportion of mothers eating more than 15 packages of food supplementation was 48% in the LFS group compared to 32% in the GFS group (p=0.225).

After 60 days treatment, mean difference between groups for MUAC was 0.18 cm (95%CI −0.25, 0.38). Mothers in the LFS group were 2.28 times more likely to increase MUAC (RR 2.28; 95%CI 1.58,3.27). The mean difference between groups for gestational weight was 0.49 kg (95%CI −0.25, 0.38). Mothers in the LFS group were 4.73 times more likely to increase weight (RR 4.73; 95%CI 1.37,16.3) than were mothers in the GFS group Please see table 5.

**Table 5.**
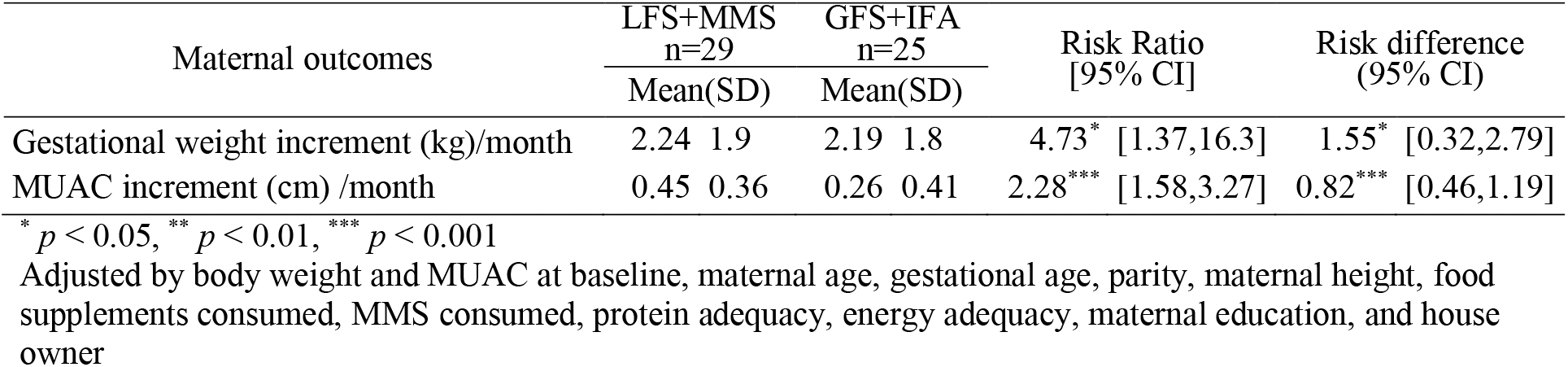
Effect of local food supplements (LFS) and MMS on maternal outcomes compare to government fortified food supplements (GFS) + IFA intervention

| Maternal outcomes | LFS+MMS<br>n=29 |  | GFS+IFA<br>n=25 |  | Risk Ratio<br>[95% CI] | Risk difference<br>(95% CI) |
| --- | --- | --- | --- | --- | --- | --- |
|  | Mean(SD) |  | Mean(SD) |  |  |  |
| Gestational weight increment (kg)/month | 2.24 | 1.9 | 2.19 | 1.8 | 4.73* [1.37,16.3] | 1.55* [0.32,2.79] |
| MUAC increment (cm) /month | 0.45 | 0.36 | 0.26 | 0.41 | 2.28*** [1.58,3.27] | 0.82*** [0.46,1.19] |

\* $p < 0.05$ , \*\* $p < 0.01$ , \*\*\* $p < 0.001$
Adjusted by body weight and MUAC at baseline, maternal age, gestational age, parity, maternal height, food supplements consumed, MMS consumed, protein adequacy, energy adequacy, maternal education, and house owner

Table 6 presents the effect of supplementation on birth outcomes. Mothers in the LFS group were less likely to have birth weight <3000grams, birth length, and cesarean delivery. Mothers who received the balance energy protein food supplementation and MMS resulted in a significant reduction of 22% in the risk of birth weight <3000grams (RR 0.15, 95% CI 0.023,0.98), reduction of 25% in the risk of short birth length (RR 0.068, 95% CI 0.005,0.93), and reduction of 32% in the risk of cesarean delivery (RR 0.11, 95% CI 0.022,0.60) respectively. There was no statistically significant effect of balanced protein energy supplements and MMS on preterm birth (RR 1.15, 95% CI 0.27,4.89).

**Table 6.**
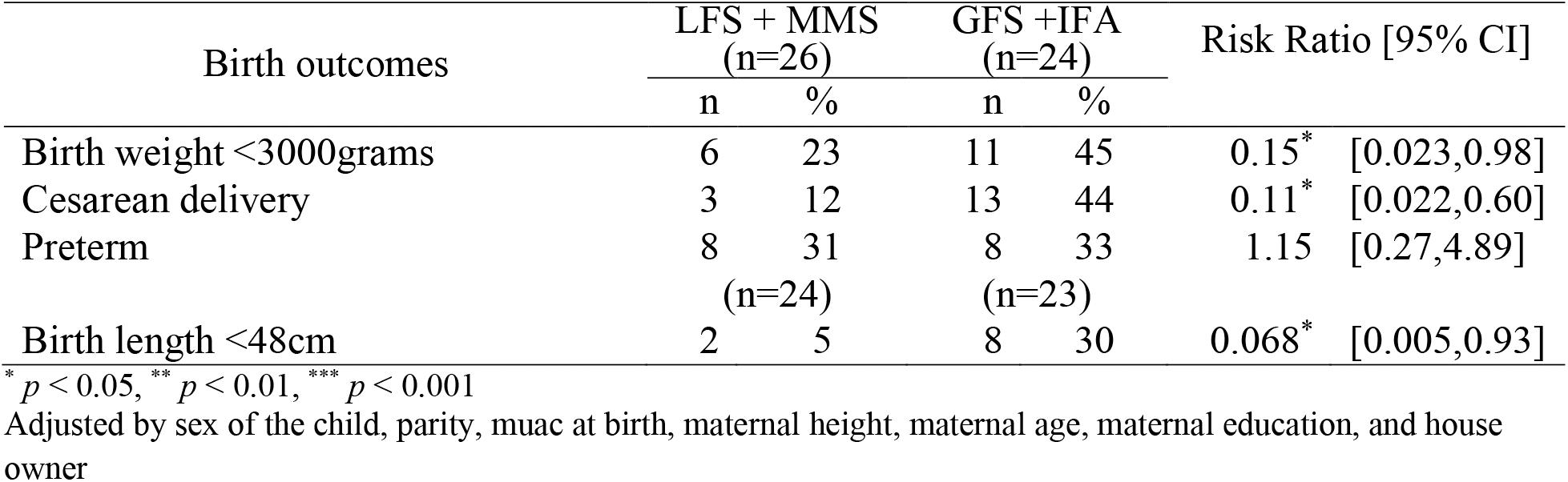
Effect of local food supplements (LFS) +MMS on birth outcomes compare to government fortified food supplements (GFS) + IFA interventions

| Birth outcomes | LFS + MMS<br>(n=26) |  | GFS +IFA<br>(n=24) |  | Risk Ratio [95% CI] |  |
| --- | --- | --- | --- | --- | --- | --- |
|  | n | % | n | % |  |  |
| Birth weight <3000grams | 6 | 23 | 11 | 45 | 0.15* | [0.023,0.98] |
| Cesarean delivery | 3 | 12 | 13 | 44 | 0.11* | [0.022,0.60] |
| Preterm | 8 | 31 | 8 | 33 | 1.15 | [0.27,4.89] |
|  | (n=24) |  | (n=23) |  |  |  |
| Birth length <48cm | 2 | 5 | 8 | 30 | 0.068* | [0.005,0.93] |

\* $p < 0.05$ , \*\* $p < 0.01$ , \*\*\* $p < 0.001$
Adjusted by sex of the child, parity, muac at birth, maternal height, maternal age, maternal education, and house owner

## Discussion

In this pilot study in rural Indonesia, the provision of low-cost local food combined with multiple micronutrient supplementation (LFS) improved indicators of maternal nutrition including gestational weight gain and improvements in MUAC. We also found significant improvements in birth weight, birth length, and decreased occurrence of cesarean delivery. However, the LFS did not impact the risk of preterm birth. We hypothesize there are three reasons for the apparent benefit of LFS including: the nutritional composition of the LFS product, the known benefits of MMS as compared to IFA alone, and the higher compliance among the LFS group.

Previous work has shown that provision of MMS, as compared to IFA alone, can improve birth outcomes. Meta-analyses on MMS showed to improve birth outcomes including reducing the risks of preterm birth, low birth weight, and small for gestational age in comparison with IFA. Furthermore, there were no adverse events for pregnant women or infants with MMS (Bourassa et al., 2019).

Previous work has shown that improving maternal intake of specific macronutrients, such as proteins, carbohydrates, and fats could improve gestational weight gain. In this study, providing food supplementation for undernourished pregnant mothers could improve weight gain and MUAC. The LFS group showed higher weight gain (2.25+1.7 kg) and MUAC increment by (0.43+0.35 cm) per month compared to the GFS group by (2.03+1.4 kg) and (0.25+0.40 cm) per month. A study in Cambodian women that providing food supplementation consumed during pregnancy did not significantly increase maternal weight gain (Janmohamed et al., 2016; Kardjati, Kusin, & Schofield, 1990). Otherwise, a study on a milk-based fortified take-home product showed that women had greater weight gain (12.29 vs 11.31 kg, p < 0.05) than those in the control group (Mardones-Santander et al., 1988).

This study shows a significant effect on birth weight and birth length. The provision of balanced energy protein food supplements and MMS could significantly 85% reduction of proportion infants who had a birth weight less than 3000 grams (OR 0.15, 95% CI 0.023,0.98) and who had short birth length <48cm (OR 0.068, 95% CI 0.005,0.93). It was in concordance with the previous study in the Gambia, which showed the odds ratio for low birthweight babies in supplemented pregnant women was 0.61 (95% CI 0.47 to 0.79, P < 0.001) (Ceesay et al., 1997). Meta-analyses from twelve studies on balance energy protein supplementation showed an improvement in mean birth weight (mean difference +40.96 g, 95%CI 4.66 to 77.26) and reduction of small for gestational age (RR 0.79, 95% CI 0.69 to 0.90) (Kramer & Kakuma, 2010; Ota, Hori, Mori, Tobe-Gai, & Farrar, 2015). This result was consistent with the study in rural Burkina Faso, which showed the clinically important of providing food fortified supplementation to underweight pregnant mothers on increased infant birth weight by 31grams and length by 4.6mm (Huybregts et al., 2009). A large cluster randomized control trial enrolled 1248 pregnant women, reported the intervention groups who were fully exposed to the lipid-based nutrition supplementations had higher length-for-age Z scores of 0·216 SD [0·043 to 0·389] and lower stunting prevalence −8·2% [−15·6 to −0·7] for T3) (Galasso, Weber, Stewart, Ratsifandrihamanana, & Fernald, 2019).

The rate of cesarean delivery in Indonesia increased almost twofold from 9.8% in 2013 to 17.6% in 2018. Perhaps surprisingly, we found that the cesarean delivery rate in the LFS treatment group was 12%, significantly much lower than 44% in the GFS control group. This suggests that balanced energy protein supplements, combined with MMS, may minimize the need for cesarean delivery. Cesarean delivery is a life-saving intervention for mothers and infants to treat complications that have short-term and long-term health effects, but overuse of unnecessary cesarean can cause harm. (Niino, 2011; Sandall et al., 2018). Unfortunately, we didn’t explore the cause of cesarean-surgical, and we could not know whether it was a medical necessity. Additionally, we found higher pregnancy complaints (e.g. nausea, headache, vomit, tired and weak, frequent urination, pelvic joint pain, lack of appetite, insomnia, back pain, fever, leucorrhoea, etc.) in participants from the GFS group than in participants from the LFS group (Susiloretni et al, unpublished research report, 2010). The decrease of the cesarean delivery rate might have benefits, especially could reduce total health expenditure by a service provider on hospital care and would promote the efficiency of the National Health Insurance System (Agustina et al., 2019). This result might support that giving BEP + MMS to the pregnant mothers did not increase cesarean delivery as some have worried.

Balanced protein energy supplementation has been shown to have significant positive impacts on the incidences of preterm birth (Villar et al., 2003). The findings from this study showed that the two treatments had similar results on average gestational age at birth (37.9 vs 37.9 weeks) and the incidences of preterm birth (31% vs 33%). There was no difference in the incidence of preterm birth between the two groups. This result supported the finding of a review article from five studies involved 3408 women, showed no significant effect of balanced energy protein supplementation for preterm birth (Ota et al., 2015). Preterm birth is the leading risk factor for infant death due to neonatal infections and contributes to long-term physical and neurological morbidity (Abubakar, Tillmann, & Banerjee, 2015; Blencowe et al., 2012; Lawn et al., 2014). The rate of preterm birth among previous chronic energy malnutrition pregnant mothers was high at above 30% almost the same with the rate national preterm birth at 29.5% (Ministry of Health Republic of Indonesia, 2018), it might be a serious concern. The government of Indonesia should give more attention to intervention to reduce the preterm birth rate. A meta-analysis showed that MMS could reduce the risks of preterm birth, low birth weight, and small for gestational age in comparison with IFA alone. As a country with inadequate micronutrient intakes, it should consider supplementing pregnant women with MMS as a cost-effective method to reduce the risk of adverse birth outcomes (Bourassa et al., 2019)

This study had many limitations. This was a small sample size study that might interfere with generalizability and insufficient to ensure balance during randomization. However, the baseline data showed no meaningful differences in key characteristics between the two groups. The compliance with the intervention was lower than expected, so we categorized the compliance for taking food and nutrient supplementations. The main reason for lower compliance was because intervention implementation; we received reports that some supplements had not been delivered to the participants. Another weakness was that 8% of women were loss to follow-up at birth follow-up and 15% at study end. The primary reason women were lost to follow up was because they moved to other areas that couldn’t be reached by data collectors.

## Conclusions for Practice

Our results support the potential for LFS, combined with MMS, to be used to improve maternal and birth outcomes as a nutrition package in a low-income setting. Low-cost local food supplementation could improve maternal and birth outcomes, namely maternal weight gain, MUAC increment, birth weight, and length, and reduced cesarean delivery. Future studies should be conducted to replicate these pilot results given the limitations of this study including a small sample size and lower than expected intervention compliance. Finally, there are important challenges to improve maternal and birth outcomes with local food supplementations, including food supplementation productions and policymaker advocacy to use evidence-based research for an intervention program. Challenges to local production may be standardizing the nutrient composition and ensuring food safety requirements. However, UNICEF and WFP have shown in recent years that local production of RUTF/RUSF products can be safely and successfully done at a large scale. Food supplementation production could empower community or medium and micro-enterprise businesses to produce. It could be income generating in a low-income setting and increase the use of local foods. Since local food supplementations adapt to sustainable development goals, these could reduce food losses and waste, shorten the food chain, reduce the effect of the environment, and improve the sustainable food system (Willett et al., 2019).

## Data Availability

Data could be obtained from the corresponding author by request.

https://clinicaltrials.gov/ct2/show/NCT02959125

---

**Supplemental file #1.** Cookies ingredients received by mothers

| **Foods** | **NutCookies 1** | **NutCookies 2** | **NutCookies 3** | **Fish snacks** |
| --- | --- | --- | --- | --- |
| Peanut | 40 g | 40 g | 40 g | - |
| Green bean | 120 g | - | - | - |
| Soybean | - | 120 g | - | - |
| Brown bean | - | - | 120 g | - |
| Wheat flour | 60 g | 60 g | 60 g | 100 g |
| Corn flour | 120 g | 120 g | 120 g | 100 g |
| Cassava flour |  |  |  | 70 g |
| Milk powder | 75 g | 75 g | 75 g |  |
| Butter | 120 g | 120 g | 120 g | 30 g |
| Sugar | 200 g | 200 g | 200 g | 45 g |
| Eggs yolk | 3 eggs | 3 eggs | 3 eggs |  |
| Eggs | - | - | - | 2 eggs |
| Fish with bone | - | - | - | 150 g |
| Purple sweet potatoes | - | - | - | 10 g |
| Vegetable oil | - | - | - | 5 g |

**Supplemental file #2.** Hedonic test for LFS and GFS: average value

| Hedonic test | LFS | GFS | p* |
| --- | --- | --- | --- |
| Flavor | 3.40 | 3.24 | 0.173 |
| Odor | 3.13 | 3.00 | 0.162 |
| Texture | 3.51 | 3.00 | 0.028 |
| Color | 3.10 | 2.96 | 0.114 |

1=very poor, 2=poor, 3=good, and 4=very good

